# Depressive symptoms are associated with differential cognitive and neuroanatomical alterations in young and older adults

**DOI:** 10.1101/2023.11.26.23299020

**Authors:** Eyal Bergmann, Daniel Harlev, Cam-CAN, Noham Wolpe

**Author notes:** Corresponding author: Send correspondence to Dr. Bergmann, Department of Psychiatry, Rambam Health Care Campus, POB 9649 Bat Galim, Haifa 31096, Israel. These authors contributed equally to this work (Eyal Bergmann, Daniel Harlev).

## Abstract

**Objective:** Depression is a heterogeneous disorder. The purpose of this article is to examine the contribution of age to this heterogeneity by characterizing the associations of depressive symptoms with cognitive performance and brain structure across the lifespan.

**Methods:** The authors analyzed demographic variables (age, gender, education), affective measures (Hospital Anxiety and Depression Scale), and cognitive assessments (The Addenbrooke’s Cognitive Examination Revised) from the Cambridge Centre for Ageing Neuroscience (Cam-CAN) cohort (N=2591, age 18-99). A subset of this cohort (N=647) underwent structural MRI, which was used for voxel-based brain morphometry.

**Results:** A linear regression model revealed a significant interaction between age and depression score, indicating that depression-related cognitive dysfunction is more severe in older adults. A comparison of different cognitive domains showed that this effect was consistent across all tested domains but significantly more prominent for fluency. A complementary voxel-based morphometry analysis, based on similar regression models, revealed age by depression interactions in several brain regions, demonstrating preferential age-related reduction in grey matter volume in the left and right hippocampi in older adults. The reciprocal contrast revealed preferential reduction in grey matter in the left superior frontal gyrus, left middle frontal gyrus, and left superior parietal lobule in younger adults.

**Conclusions:** These findings indicate that the associations of depression with cognitive performance and brain structure are age-dependent, suggesting that the neuropathological mechanisms underlying depression may differ between young and older adults. Recognizing these differences will support the development of better diagnostic tools and therapeutic interventions for depression across the lifespan.

## Introduction

Depression is a complex and heterogeneous psychiatric disorder that affects many individuals across the lifespan (1). The definition of clinical depression, or Major Depressive Disorder (MDD), is based on categorical diagnoses as outlined in diagnostic manuals, such as the ICD-11 and DSM 5 (2, 3). However, these diagnostic categories often fall short of fully characterizing each individual’s unique clinical profile and thus fail to capture the significant heterogeneity of the disorder. This is important clinically, because failing to distinguish between neurobiologically distinct clinical entities within depression (4) can lead to ineffective treatment (5). A major goal in psychiatric research is thus to better characterize patient variability to support better diagnosis of depression subtypes and treatment personalization.

A major contributor to the clinical heterogeneity of depression is variability in symptoms. For example, the criteria for MDD typically encompasses a wide spectrum of various, at times contradictory, symptomatology (6), such as insomnia and hypersomnia. Moreover, across symptoms, patients may present with very different symptomatology, but still be diagnosed with depression. In the extreme case, two patients diagnosed with MDD may share no single symptom, Symptom heterogeneity may also be expressed in symptom severity, which can lead to ‘subclinical’ depressive symptoms that fall below the diagnostic threshold for MDD (7). Studies adopting such a ‘dimensional psychiatry’ approach for depression (8) have shown that such depressive symptoms still have a substantial impact on well-being and psychosocial functioning (9), emphasizing the need for more patient-specific intervention strategies.

Another major source of heterogeneity in depression is sociodemographic variation, and particularly aging. When compared to Early Life Depression (ELD), Late Life Depression (LLD) is often associated with poorer treatment response (10), as well as with distinct symptomatology (11) and brain alterations (12). Moreover, multiple cognitive domains are impacted in depression, and the specific domains impacted by depression may differ in young vs. older adults, although conflicting evidence has been reported (13) .

The diverse symptomatology, brain changes, and cognitive impairments in ELD and LLD suggest that these conditions, although diagnosed and treated similarly, may reflect distinct pathophysiology. However, to date, studies have predominantly focused on either young or elderly populations separately, often failing to capture the complex interaction between normative aging processes, depressive symptoms and consideration of specific cognitive changes. In addition, most studies define depression as a categorical disorder rather than a dimensional condition (8). Here, we sought to investigate the interaction between age, depressive symptoms and cognitive functions in a large, population-derived cohort of individuals across the lifespan. We examined depressive symptoms along a continuum of severity, while accounting for medication status, and compared the cognitive and neuroanatomical differences associated with depressive symptoms between young and older adults. Our overarching hypothesis is that depressive symptoms in young and older adults represent distinct clinical entities. More specifically, we hypothesized that depressive symptoms would be associated with distinct cognitive and brain differences in older vs. young adults.

## Methods

### Participants

We conducted data analysis utilizing the extensive dataset from the Cambridge Centre for Ageing and Neuroscience (Cam-CAN), which is a large-scale project focusing on normative aging (14). The initial recruitment process involved an opt-out strategy to maximize representation across different population groups, with a similar representation across age deciles, as previously described (15). Data included in this study was from Stage 1 (CC3000) and Stage 2 (CC700). Ethical approval was granted by the local ethics committee, Cambridgeshire 2 (now East of England – Cambridge Central) Research Ethics Committee (reference: 10/H0308/50). Written informed consent was obtained from all participants before commencing the study. The study conforms to the provisions of the Declaration of Helsinki.

### Behavioral data acquisition

In Stage 1, n=2681 participants completed a home interview, providing detailed demographic questionnaires on age, sex, education level, lifestyle, and medical history. Cognitive functioning was assessed using various neuropsychological tests, including the Addenbrooke’s Cognitive Examination-Revised (16), which is a widely used cognitive battery designed for dementia screening that assesses multiple cognitive domains, namely orientation, memory, fluency, language, and visuospatial abilities. As the domains differ in their maximal score, the values were normalized to a range between 0 and 1 for interpretability. Affective symptoms were measured using the Hospital Anxiety and Depression Scale (17), which is a 14-item self-reported questionnaire with depression (HADS-D) and anxiety (HADS-A) subscales. After exclusion of participants with missing demographic or clinical information, the final cohort included 2591 participants. **Table 1** provides an overview of the key variables, including age, sex, scores on the HADS-D, HADS-A, and the ACE-R, across the age deciles.

**Table 1.** demographic, affective and cognitive variables in Cam-CAN cohort.

| Age | N | Sex<br>M/F | HADS-D<br>mean (SD) | HADS-A<br>mean (SD) | ACE-R<br>mean (SD) | Education* |  |  |  |
| --- | --- | --- | --- | --- | --- | --- | --- | --- | --- |
|  |  |  |  |  |  | None | GCSE | A Levels | University |
| 18-29 | 248 | 103 / 145 | 3.39 (2.73) | 6.17 (3.56) | 93.2 (6.92) | 4 | 14 | 45 | 185 |
| 30-39 | 354 | 153 / 201 | 2.89 (3.11) | 5.98 (3.32) | 94.1 (6.33) | 5 | 8 | 14 | 327 |
| 40-49 | 314 | 138 / 176 | 2.78 (2.75) | 5.46 (3.26) | 94.8 (5.36) | 10 | 15 | 9 | 280 |
| 50-59 | 263 | 128 / 135 | 2.85 (3.22) | 5.72 (3.68) | 93.8 (6.46) | 19 | 9 | 17 | 218 |
| 60-69 | 375 | 175 / 200 | 3.02 (2.74) | 4.91 (3.18) | 93.3 (6.75) | 54 | 34 | 17 | 270 |
| 70-79 | 434 | 207 / 227 | 3.01 (3.33) | 4.77 (3.39) | 89.4 (8.07) | 119 | 33 | 16 | 266 |
| 80-89 | 521 | 210 / 311 | 4.24 (2.92) | 4.31 (3.14) | 84.6 (10.8) | 173 | 35 | 17 | 296 |
| 90-99 | 82 | 23 / 59 | 4.84 (3.14) | 4.22 (3.26) | 79.7 (11.5) | 35 | 10 | 0 | 37 |
| Total | 2591 | 1137 / 1454 | 3.31 (2.9) | 5.16 (3.39) | 90.8 (8.97) | 419 | 158 | 135 | 1879 |

\* Categorized according to the British education system: “none” = no education over the age of 16 y; “GCSE” = General Certificate of Secondary Education; “A Levels” = General Certificate of Education Advanced Level; “University” = undergraduate or graduate degree.

### Structural neuroimaging acquisition and pre-processing

In Stage 2, a subset of participants (n=656) underwent more extensive cognitive testing and neuroimaging. Exclusion criteria are described at length in the original report (14) including: significant cognitive impairment, communication difficulties, significant medical problems, mobility problems, substance abuse, and MRI/MEG safety and comfort issues.

Neuroimaging data acquisition and pre-processing is described at length in Taylor et al. (15). In brief, participants were scanned with a 3T Siemens TIM Trio with a 32-channel head coil. A T1-weighted MPRAGE image was acquired (repetition time 2250 ms, echo time 2.99 ms, inversion time 900 ms, field angle 9°, field-of-view 256 mm × 240 mm × 192 mm, isotropic 1 mm voxels.) The MRI data of six participants were not included in the analysis due to technical problems during scanning or pre-processing. Together with the exclusion of three participants due to missing sociodemographic information, 647 participants were included in the structural imaging analyses. A comparison between the clinical and demographic data of participants included in stage 1 and stage 2 is shown in **Suppl. Table 1.** The structural images were pre-processed for a voxel-based morphometry (VBM) analysis using SPM12 (www.fil.ion.ucl.ac.uk/spm) as previously described (15). Multimodal segmentation was used to reduce age-biased tissue priors. Diffeomorphic Anatomical Registration Through Exponentiated Lie Algebra approach was applied to improve inter-participant alignment (18) as follows. Segmented images were warped to a project-specific template while modelling the shape of each brain. The resulting images were affine transformed to the Montreal Neurological Institute space using the template and individual brain parameters. Voxel size of the normalized images was 1 mm isotropic. These normalization steps were followed by modulation by the Jacobean of the combined transformations (to preserve volume) and smoothing with an 10-mm full width at half maximum (19).

### Statistical analysis

Statistical analysis of behavior examined the relationship between depressive symptoms and cognitive performance. Descriptive statistics and regression models were employed to explore the associations of interest: to examine the effects of depression and age on total ACE-R score, we used a linear regression model with HADS-D, age and their interaction as covariates. Sex (categorical variable), education (ordinal variables), current use of antidepressants (categorical variable), anxiety (HADS-A), and anxiety by age interaction were included as covariates of no interest. All analyses were performed with R version 2023.09.0+463 (20). For the comparison between cognitive domains, a liner-mixed effects model was used using *lmer* package (21). The interaction of cognitive domain, age and depression was calculated using type III analysis of variance (22). Bayesian Information Criteria was used to compare model goodness of fit (23).

For the structural imaging analyses, a multiple regression was conducted to generate a statistical parametric map of differences in grey matter volume in relation to age and depression. Depression, age, and the (mean-corrected and orthogonalized) interaction term between depression and age were included as the main covariates of interest. Covariates of no interest included those from the regression model of behaviour (above), in addition to handedness as a numerical variable (24) and total intracranial volume. Clusters were identified at *p* < 0.05, family-wise-error-(FWE-) corrected. Labelling of significant clusters was down using MRIcron software (https://people.cas.sc.edu/rorden/mricron/). For visualization purposes in the figures of interactions in both behavioural and neuroimaging data, participant data were grouped into categorical age groups (18-39, 40-64, 65+) and depression (HADS-D<4, HADS-D=4-7, HADS-D>7) groups (17). However, all analyses were performed with age and depression as continuous variables.

## Results

### Depression and cognition across age

To characterize the relationship between depression and cognition across age, we submitted these measures to a linear regression analysis. The model was significant (Adjusted R² = 0.279, P < 0.001), and showed that both HADS-D and HADS-A were significantly associated with ACE-R scores (**Supp. Table 2**), such that higher HADS-D but lower HADS-A, were associated with lower ACE-R. Importantly, including age x HADS-D and age x HADS-A interactions in the model significantly improved the model fit (R²=0.289, BIC of 17912 vs. 17930). This model (**Table 2**) showed a positive age x HADS-D interaction, with an age-dependent increase in the effect of depression (**Fig. 1A**). In addition, a negative age x HADS-A interaction was observed with an age-dependent decrease in the effect of anxiety on ACE-R. Collectively, these results suggest differential age-dependent relationship between affective symptoms and global cognitive performance. We next examined the association between these affective symptoms and specific cognitive domains.

**Figure 1.**
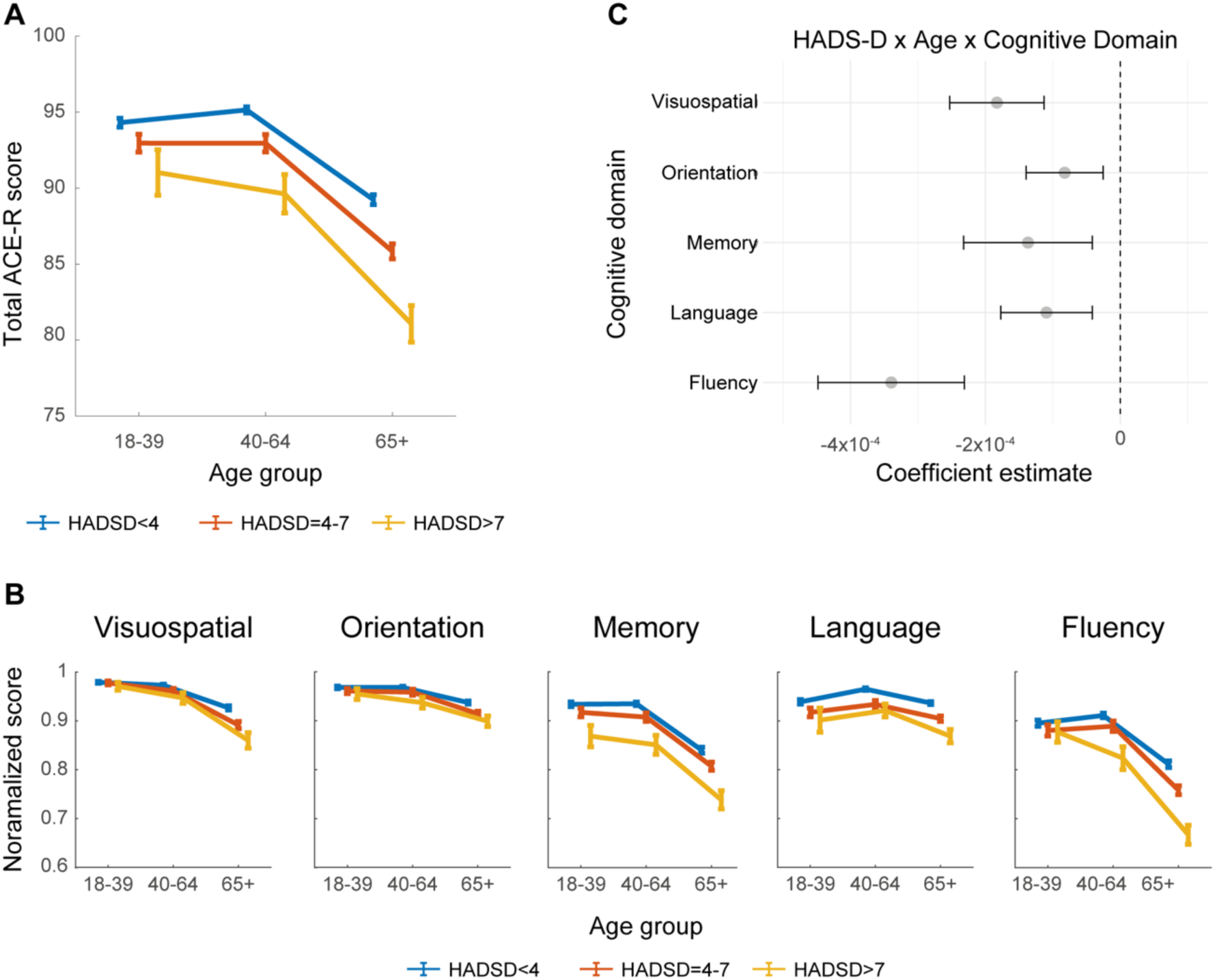
The association between age, depression, and cognitive performance. (A) Illustration of the significant interaction effect of age x depression on total ACE-R score. Total ACE-R score plotted against age for different HADS-D scores. Error bars represent the standard error of the mean. Age and depression scores are grouped for visualization purposes only and were entered as continuous variables into the regression models. (B) Same as (A), but for the specific ACE-R domains. As the domains differ in their maximal score, the values were normalized to range between 0 and 1 for interpretability. Error bars represent the standard error of the mean. (C) Illustration of the effects of age × depression interaction on specific cognitive domains. Grey dot indicates mean beta coefficient of the interaction term for each domain, with error bars showing 95% confidence interval.

**Table 2.** Demographic and affective predictors of total ACE-R score in the linear regression model.

| Total ACE-R score |  |  |  |  |
| --- | --- | --- | --- | --- |
| Variable | $\beta$ estimate | $\beta$ SE | z-value | p-value |
| (Intercept) | 93.26 | 1.05 | 88.95 | < **0.001** |
| Sex | -0.10 | 0.01 | -7.13 | 0.82 |
| Education | -0.07 | 0.30 | -0.23 | < **0.001** |
| Antidepressants | 2.33 | 0.14 | 16.76 | **0.034** |
| Age | -1.37 | 0.64 | -2.13 | < **0.001** |
| HADS-D | 0.33 | 0.19 | 1.79 | 0.073 |
| HADS-A | -0.43 | 0.15 | -2.80 | **0.005** |
| Age × HADS-D | -0.02 | 0.00 | -5.57 | < **0.001** |
| Age × HADS-A | 0.01 | 0.00 | 3.89 | < **0.001** |

A linear mixed model revealed as significant interactions between cognitive domain, age, and HADS-D (Wald χ2(4) = 36.4, P < 0.001) and cognitive domain, age and HADS-A (Wald χ2(4) = 17.39, P = 0.002). To break down this 3-way interaction, we repeated the analysis for each domain separately and compared the estimates of age x HADS-D interactions between each pair of cognitive domains (**Fig. 1B**). While all domains showed a significant association with the interaction between age x HADS-D, the magnitude of these associations differed (**Fig. 1C, Suppl. Table 3-6**). A statistical comparison between domains showed stronger interaction in fluency, compared to orientation (t = 4.73, P < 0.001), memory (t = 3.96, P < 0.001) and language (t = 4.61, P < 0.001). A marginally significant difference was also observed between fluency and visuospatial abilities (t = 2.67, P = 0.075, all tests were Bonferroni corrected for multiple comparisons). Comparisons between other pairs of domains were insignificant (**Suppl. Table 7**). Together, these findings indicate a reduction in cognitive performance with increased depressive symptoms and age, with fluency showing the most significant reduction across most cognitive domains.

### Depression and brain structure across age

To characterize the brain correlates of depressive symptoms by age, we used VBM in 647 participants that underwent brain imaging. This subset of participants was more balanced in terms of age, sex distribution and level of education. Importantly, they all had lower HADS-D and higher ACE-R scores compare to subset of participants that were not scanned (**Suppl. Table 1**).

The structural brain imaging analyses showed five clusters where grey matter volume was associated with age x HADS-D, i.e., with age-dependent effects of depression (**Fig. 2A**, **Table 3**). To interpret this interaction, we plotted grey matter volume from these areas as a function of age and HADS-D score (**Fig. 2B**). There was an age-dependent increase in the relationship between HADS-D and brain volume in the left and right hippocampi. By contrast, the left superior frontal gyrus (SFG), middle frontal gyrus (MFG) and left superior parietal lobule (SPL) showed an age-dependent decrease in this relationship. Collectively, these results indicate that depressive symptoms are associated with different brain structures as a function of age.

**Table 3:** Summary of peak coordinates and statistics for the cluster showing an age x HADS-D interaction. KE indicates cluster size in number of voxels.

| Brain region |  | K<sub>E</sub> | Coordinates |  |  | T-score |
| --- | --- | --- | --- | --- | --- | --- |
|  |  |  | x | y | z |  |
| Positive interaction |  |  |  |  |  |  |
| L | Superior Frontal Gyrus | 516 | -21 | 44 | 47 | 5.3 |
| L | Middle Frontal Gyrus | 511 | -50 | 32 | 31 | 5.28 |
| L | Lateral Occipital Cortex | 380 | -27 | -62 | 66 | 5.54 |
|  |  |  | -16 | -70 | 64 | 4.58 |
| Negative interaction |  |  |  |  |  |  |
| L | Hippocampus | 898 | -27 | -12 | -13 | 5.02 |
| R | Hippocampus | 214 | 25 | -13 | -11 | 4.64 |

**Figure 2.**
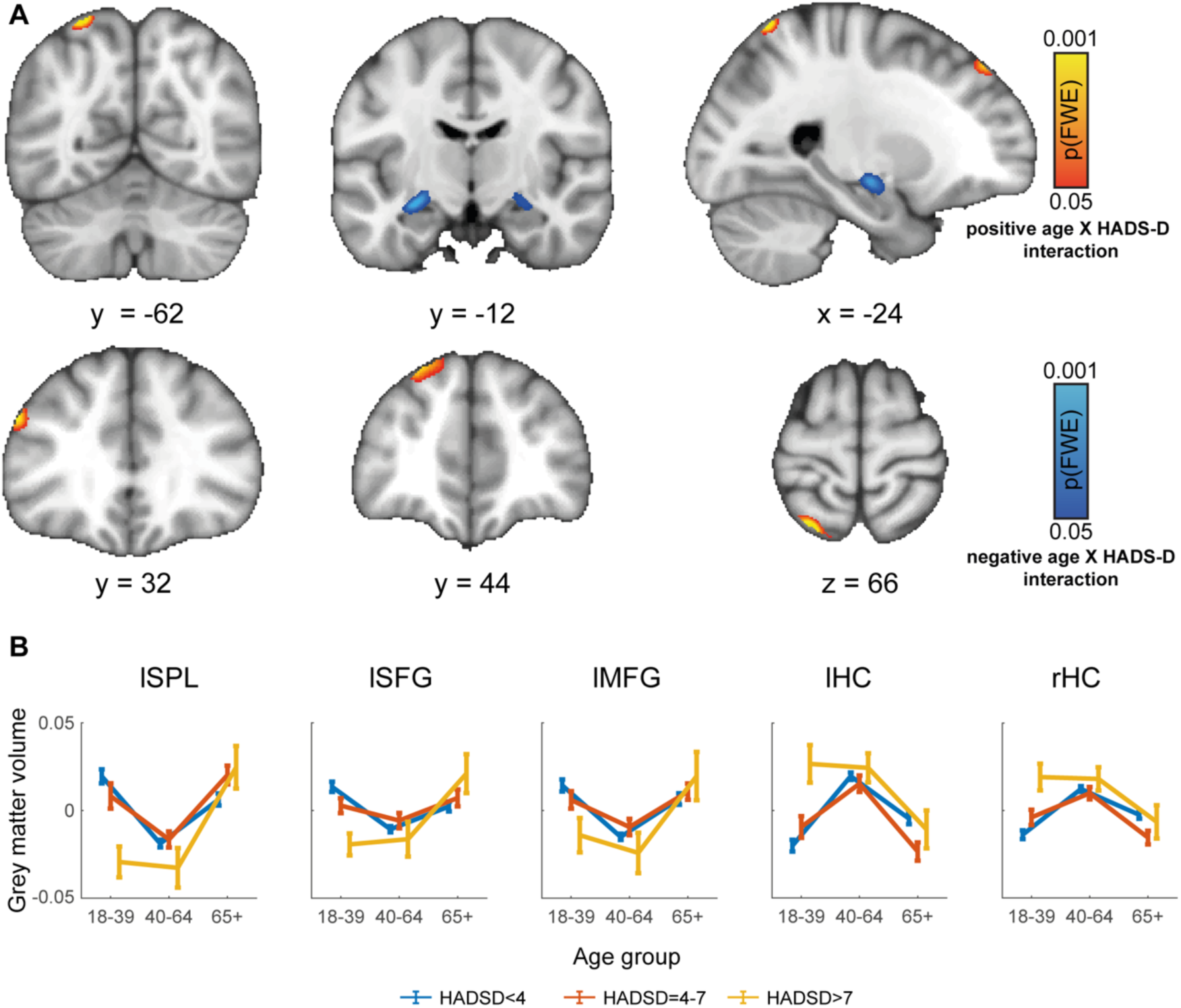
Age-dependent differences in structural correlates of depressive symptoms. (A) Voxel-based morphometry results showing significant interactions between age and HADS-D. (B) Illustration of the effects of age and depression on grey matter volume from (A). The results illustrate the differences grey matter volume (mean corrected) across age, as a function of HADS-D score. Results are shown for the five clusters from (A), labelled according to MRIcron. Note that HADS-D and age are displayed as categorical variables for illustration purposes but were entered as continuous variables in the analyses.

## Discussion

Our findings show that the association between depression and reduced cognitive performance is more pronounced with aging. This was found across different cognitive domains and was particularly prominent for fluency. Moreover, structural brain imaging revealed that depressive symptoms are associated with reduced grey matter volume in different brain regions as a function of age. Together, our findings suggest that the associations of depression with cognition and brain structure are different for young and older adults. These results emphasize the possibly distinct pathophysiology underlying depression across the lifespan.

Previous studies examined the effect of ELD and LLD on cognition separately. These studies reported a modest reduction in executive functions and memory in ELD (25). In LLD, a reduction in cognitive performance was found in multiple domains, including attention, executive functions, memory, and processing speed (26). Among these domains, reduced cognitive performance in LLD was most pronounced in executive functions (27), with approximately 30% of depressed older adults exhibiting impaired performance in this domain (28). Here, we replicated a depression-related reduction in cognitive performance with age, which was most pronounced in the fluency domain – a cognitive domain strongly dependent on executive control (29). Importantly, by analyzing these effects continuously across the lifespan within the same study, we showed that this effect is age-specific, as it was not observed in younger adults, which is consistent with a modest depression-related cognitive changes in young adults (30).

Our finding that depressive symptoms in old age are associated with reduced executive function is in close agreement with research on the so-called ‘Depression-Executive Dysfunction’ (DED) syndrome. This clinical syndrome presents a unique combination of neurocognitive and neuropsychiatric features (31). DED syndrome is characterized by a combination of neurocognitive deficits, alongside neuropsychiatric symptoms such as apathy and psychomotor retardation (32). These features collectively contribute to a distinct clinical presentation in older adults with LLD. The clinical significance of the DED syndrome is substantial, as executive dysfunction has been identified as a predictor of poor response to antidepressant treatments in late-life depression (31). Additionally, it is associated with early relapse and recurrence of depression in old age, highlighting the importance of considering the cognitive dimension of depression in older adults when designing treatment strategies (28).

The analysis of brain structure identified several brain regions that are differentially affected by depression across the lifespan. In older adults, depressive symptoms were associated with reduced hippocampal volume compared to their non-depressed counterparts, whereas in young adults the opposite trend was observed. This finding is consistent with a recent study pointing to a similar link in subclinical depressive symptoms in older adults (33). Importantly, the age-dependent effect explains the variability reported in previous studies of MDD, as differences in hippocampal volumes were observed in older, but not young cohorts (34). Conversely, in younger adults, we found that depressive symptoms are associated with reduced grey matter volume in frontal and parietal association cortices. These regions are typically part of the Default, Dorsal Attention and Frontoparietal networks – networks which typically show alterations in MDD as found in a recent meta-analysis (35). Examining age distributions in this meta-analysis (c.f. eTable 1), the average age was 37.77 years with only 20% of the participants being older than 65. Another meta-analysis by Zhukovsky et al. compared structural and functional alterations in ELD with MDD and LLD (36). The results revealed that changes in frontoparietal and dorsal attention networks were common to both age groups. Our findings are somewhat inconsistent, we found preferential reduction in grey matter volume in young adults with depressive symptoms in regions of these networks. This may stem from different neuropathology underlying subclinical depression and MDD, but a more likely explanation is age differences in the LLD group that was examined. In the 17 studies included in Zhukovsky et al., the mean age was 68.4 years (SD = 6.1 years, range 55-79 years), and the total number of ELD cases was much larger the LLD (6362 vs. 535). In contrast, in our study more than a third of the participants in the neuroimaging cohort were over 65 and were recruited using the same approach as the young participants, reducing the risk for a selection bias. Yet, further research is needed to address this question, and a promising avenue is longitudinal imaging of participant over aging to examine within-participant effects.

The distinct cognitive and neuroanatomical correlates of depressive symptoms found across age support the possibility that depression in young and older adults reflects a distinct clinical condition (37). This has important clinical implications, as most clinical trials of depression treatment have been conducted in young adults, and, indeed, depressed older adults show poorer clinical response to these treatments (10). Moreover, our study is consistent with previous research showing that in LLD, depression may be both a risk factor for, and a manifestation of cognitive decline (38). A better understanding of depression heterogeneity in general, and with aging in particular, offers promising prospects for more tailored interventions in the future.

### Strengths and limitations

The presented research benefits from several noteworthy strengths. The study employs validated scales and cognitive assessment tools with robust structural imaging analyses in a large population-derived cohort across the lifespan. Participants were diverse and more representative of the population, with balanced sociodemographic characteristics, spanning all age groups and with a similar number of participants in each age decile. All analyses were performed with age, affective symptoms, cognitive performance, and brain structure measures as continuous variables, which takes into account the heterogeneity in population. These features improve generalizability, which is so urgently needed in clinical research. However, it is important to acknowledge the limitations of our study. First, its cross-sectional nature hinders our ability to establish causal relationships or evaluate alterations in depressive symptoms and neuroimaging patterns over time. Second, in the current study, we looked at depressive symptoms without accounting for symptom identity, limiting inferences about the contribution of specific symptoms to cognitive and brain structure measures. Third, the assessment of depressive symptoms relied on self-report questionnaires, which are inherently subjective and may be influenced by factors such as the participant’s honesty, self-awareness, and introspective abilities.

## Conclusion

Our study reveals that while depression and aging are consistently associated with cognitive deficits, cognitive domains are differentially affected by depression as a function of age, with fluency being particularly affected with depressive symptoms in older adults. Moreover, the distinct brain correlates of depression across age in our study raise the possibility for different underlying biological underpinnings of this complex heterogenous disorder across the lifespan. Further research is warranted to explore these dynamics in greater detail, potentially paving the way for more targeted diagnostic markers and therapeutic interventions tailored to specific age groups.

## Data Availability

All data produced in the present study are available upon reasonable request at Cam-CAN (Cambridge Centre for Ageing Neuroscience) dataset inventory.

https://camcan-archive.mrc-cbu.cam.ac.uk/dataaccess/

## Acknowledgements

We are grateful to the Cam-CAN respondents and their primary care teams in Cambridge for their participation in this study. We thank Dr. Ramit Ravona-Springer for her helpful comments on this work.

## Supplementary Material

**Supplementary Table 1.** Demographic, affective, and cognitive variables in participant subgroups with and without MRI data.

| Variables | Number (percentage %) or mean (standard deviation) |  |  |
| --- | --- | --- | --- |
|  | MRI+ (n=647) | MRI- (n=1944) | p-value |
| Age | 54.7 (18.6) | 62.2 (21.2) | p < 0.001 |
| Sex (=female) | 328 (50.7%) | 1126(57.9%) | p < 0.001 |
| Education |  |  | p < 0.001 |
| None | 49 (7.6%) | 370 (19%) |  |
| GCSE | 43 (6.6%) | 115 (5.9%) |  |
| A levels | 36 (5.6%) | 99 (5.1%) |  |
| University | 519 (80.2%) | 1360 (70%) |  |
| HADS-D | 2.77 (2.54) | 3.49 (2.99) | p = 0.001 |
| HADS-A | 4.97 (3.31) | 5.22 (3.42) | p < 0.001 |
| ACE-R | 94.9 (4.71) | 89.4 (9.61) | p = 0.95 |

**Supplementary Table 2.** First order demographic and affective predictors of cognition (total ACE-R score).

| Predictors | $\beta$ estimate | $\beta$ SE | z-value | p-value |
| --- | --- | --- | --- | --- |
| (Intercept) | 93.02 | 0.77 | 120.16 | **<0.001** |
| Age | -0.11 | 0.01 | -13.00 | **<0.001** |
| Sex | 0.06 | 0.31 | 0.20 | 0.843 |
| Education | 2.38 | 0.14 | 16.98 | **<0.001** |
| Antidepressants use | -1.21 | 0.65 | -1.87 | 0.062 |
| HADS-D | -0.65 | 0.06 | -10.62 | **<0.001** |
| HADS-A | 0.15 | 0.04 – 0.25 | 2.78 | **0.006** |

**Supplementary Table 3.** Demographic and affective predictors of orientation.

| Orientation |  |  |  |  |  |
| --- | --- | --- | --- | --- | --- |
| Predictors | $\beta$ estimate | $\beta$ SE | z-value | p-value | |
| (Intercept) | 0.95 | 0.01 | 86.66 | <0.001 |  |
| Age | 0.00 | 0.00 | -3.16 | 0.002 |  |
| Sex | 0.01 | 0.00 | 2.68 | 0.007 |  |
| Education | 0.02 | 0.00 | 10.83 | <0.001 |  |
| Antidepressants use | -0.01 | 0.01 | -1.62 | 0.105 |  |
| HADS-D | 0.00 | 0.00 | 1.10 | 0.272 |  |
| HADS-A | 0.00 | 0.00 | -1.34 | 0.181 |  |
| Age × HADS-D | 0.00 | 0.00 | -2.84 | 0.005 | |
| Age × HADS-A | 0.00 | 0.00 | 1.43 | 0.152 | |
| R<sup>2</sup> / R<sup>2</sup> adjusted |  | 0.119 |  |  |  |

**Supplementary Table 4.** Demographic and affective predictors of memory.

| Memory |  |  |  |  |  |
| --- | --- | --- | --- | --- | --- |
| *Predictors* | $\beta$ estimate | $\beta$ SE | z-value | p-value | |
| (Intercept) | 0.95 | 0.02 | 52.26 | <**0.001** |  |
| Age | 0.00 | 0.00 | -8.11 | <**0.001** |  |
| Sex | -0.02 | 0.01 | -4.04 | <**0.001** |  |
| Education | 0.03 | 0.00 | 12.74 | <**0.001** |  |
| Antidepressants use | -0.03 | 0.01 | -2.70 | **0.007** |  |
| HADS-D | 0.00 | 0.00 | 0.34 | 0.735 |  |
| HADS-A | -0.01 | 0.00 | -2.15 | **0.032** |  |
| Age × HADS-D | 0.00 | 0.00 | -2.81 | **0.005** | |
| Age × HADS-A | 0.00 | 0.00 | 2.89 | **0.004** | |
| $R^2$ / $R^2$ adjusted | | 0.221 / 0.219 | | | |

**Supplementary Table 4a.** Demographic and affective predictors of fluency.

| Fluency |  |  |  |  |  |
| --- | --- | --- | --- | --- | --- |
| Predictors | $\beta$ estimate | $\beta$ SE | z-value | p-value | |
| (Intercept) | 0.89 | 0.02 | 42.71 | <0.001 |  |
| Age | 0.00 | 0.00 | -5.90 | <0.001 |  |
| Sex | -0.01 | 0.01 | -1.39 | 0.165 |  |
| Education | 0.04 | 0.00 | 12.73 | <0.001 |  |
| Antidepressants use | -0.01 | 0.01 | -0.70 | 0.486 |  |
| HADS-D | 0.01 | 0.00 | 2.71 | 0.007 |  |
| HADS-A | -0.01 | 0.00 | -2.65 | 0.008 |  |
| Age × HADS-D | 0.00 | 0.00 | -6.14 | <0.001 | |
| Age × HADS-A | 0.00 | 0.00 | 3.99 | <0.001 | |
| $R^2$ / $R^2$ adjusted | | 0.221 / 0.219 | | | |

**Supplementary Table 5.** Demographic and affective predictors of language.

| **Language** |  |  |  |  |
| --- | --- | --- | --- | --- |
| Predictors | $\beta$ estimate | $\beta$ SE | z-value | p-value |
| (Intercept) | 0.91 | 0.01 | 70.35 | **<0.001** |
| Age | 0.00 | 0.00 | -0.54 | 0.588 |
| Sex | 0.01 | 0.00 | 2.50 | **0.013** |
| Education | 0.02 | 0.00 | 9.41 | **<0.001** |
| Antidepressants use | 0.00 | 0.01 | 0.10 | 0.917 |
| HADS-D | 0.00 | 0.00 | 0.12 | 0.905 |
| HADS-A | 0.00 | 0.00 | -1.48 | 0.138 |
| Age × HADS-D | 0.00 | 0.00 | -3.16 | **0.002** |
| Age × HADS-A | 0.00 | 0.00 | 2.31 | **0.021** |
| $R^2$ / $R^2$ adjusted | | 0.091 / 0.088 | | |

**Supplementary Table 6.** Demographic and affective predictors of visuospatial abilities.

| Predictors | $\beta$ estimate | $\beta$ SE | z-value | p-value |
| --- | --- | --- | --- | --- |
| (Intercept) | 0.96 | 0.01 | 71.54 | **<0.001** |
| Age | 0.00 | 0.00 | -5.15 | **<0.001** |
| Sex | 0.01 | 0.00 | 3.30 | **0.001** |
| Education | 0.02 | 0.00 | 11.78 | **<0.001** |
| Antidepressants use | -0.02 | 0.01 | -2.16 | **0.031** |
| HADS-D | 0.01 | 0.00 | 3.16 | **0.002** |
| HADS-A | 0.00 | 0.00 | -1.81 | 0.070 |
| Age × HADS-D | 0.00 | 0.00 | -5.13 | **<0.001** |
| Age × HADS-A | 0.00 | 0.00 | 2.24 | **0.025** |
| $R^2$ / $R^2$ adjusted | 0.188 / 0.185 | | | |

**Supplementary Table 7.** Comparison of age-related interaction with depression across cognitive domains.

| Domain 1 | Domain 2 | t-value | Uncorrected p-value* |
| --- | --- | --- | --- |
| Fluency | Orientation | 4.73 | **< 0.001** |
| Fluency | Language | 4.61 | **< 0.001** |
| Fluency | Memory | 3.96 | **< 0.001** |
| Fluency | Visuospatial | 2.67 | 0.008 |
| Visuospatial | Orientation | 2.64 | 0.008 |
| Visuospatial | Language | 2.02 | 0.043 |
| Visuospatial | Memory | 1.17 | 0.242 |
| Memory | Orientation | 0.91 | 0.36 |
| Memory | Language | 0.32 | 0.752 |
| Language | Orientation | 0.75 | 0.464 |

\* The significance level (alpha) was adjusted for multiple comparisons using the Bonferroni correction. The original alpha level of 0.05 was divided by the number of comparisons conducted (10), resulting in a Bonferroni-adjusted alpha of 0.005. Statistical significance was determined using this adjusted alpha level.

